# Distinct Cytokine and Cytokine Receptor Expression Patterns Characterize Different Forms of Myositis

**DOI:** 10.1101/2025.02.17.25321047

**Authors:** Raphael A. Kirou, Iago Pinal-Fernandez, Maria Casal-Dominguez, Katherine Pak, Corinna Preusse, Dilbe Dari, Stefania Del Orso, Faiza Naz, Shamima Islam, Gustavo Gutierrez-Cruz, Elie Naddaf, Teerin Liewluck, Werner Stenzel, Albert Selva-O’Callaghan, Jose C. Milisenda, Andrew L. Mammen

## Abstract

**Objective:** Myositis is a heterogeneous family of inflammatory myopathies. We sought to define the differential expression of cytokines, cytokine receptors, and immune checkpoint genes in muscle biopsies from patients with different forms of myositis in order to characterize patterns of inflammation in each.

**Methods:** Bulk RNA sequencing was performed on muscle biopsy samples from 669 patients, including 105 with dermatomyositis, 80 with immune-mediated necrotizing myopathy (IMNM), 65 with anti-synthetase syndrome, 53 with inclusion body myositis (IBM), 19 with anti-PM/Scl myositis, 310 with other inflammatory or genetic myopathies, and 37 controls with normal tissue (NT). Myositis clinical groups and autoantibody subgroups were analyzed separately. Expression data was analyzed for 338 genes encoding cytokines, cytokine receptors, and immune checkpoints. Myositis group-specific genes were identified from this list by finding genes that were specifically differentially expressed in one group compared to all samples and compared to NT (α<0.001).

**Results:** IBM patients had the most differentially overexpressed genes (71) among all clinical groups, including 37 that were IBM-specific. Among the top genes were several involved in type 1 inflammation, including *CCL5*, *CXCR3*, *CCR5*, *CXCL9*, and *IFNG*. Anti-Jo1 and anti-PM/Scl patients exhibited differential overexpression of a similar set of genes, while dermatomyositis patients exhibited differential overexpression of a different set of genes involved in type 1 inflammation. IMNM patients had the least number of differentially overexpressed genes with no predominant inflammatory pattern.

**Conclusion:** Each myositis clinical group and autoantibody subgroup had differentially overexpressed inflammatory mediators, including a strong type 1 inflammatory gene signature in IBM.

**Key Messages:**

- Inclusion Body Myositis (IBM) muscle biopsies exhibit differential overexpression of a set of genes involved in type 1 inflammation.
- The *CCL5*-*CCR5* and *XCL1*-*XCL2*-*XCR1* axes are specifically differentially overexpressed in IBM muscle and may contribute to T_C_1-mediated inflammation.
- Dermatomyositis, anti-Jo1 myositis, and anti-PM/Scl myositis muscle biopsies also exhibit overexpression of type 1 inflammatory genes, but to a lesser extent than IBM.

## INTRODUCTION

Myositis refers to several major groups of inflammatory diseases of the skeletal muscle, including dermatomyositis (DM), immune-mediated necrotizing myopathy (IMNM), anti-synthetase syndrome (ASyS), inclusion body myositis (IBM), and overlap myositis, among others. Each of these clinical groups can be further divided into autoantibody subgroups, based on seropositivity for myositis autoantibodies. These include anti-Mi2 DM (Mi2), anti-MDA5 DM (MDA5), anti-NXP2 DM (NXP2), anti-TIF1γ DM (TIF1), anti-HMGCR IMNM (HMGCR), anti-SRP IMNM (SRP), anti-Jo1 ASyS (Jo1), and anti-PM/Scl overlap myositis (PM/Scl). Clinicopathologically-defined myositis groups and autoantibody-defined subgroups are characterized by distinct clinical phenotypes and muscle biopsy features with unique transcriptomic signatures[1, 2].

Autoimmune diseases are frequently characterized by specific patterns of inflammation, including type I interferon (IFN-I)-mediated inflammation, type 1 (T helper 1-mediated) inflammation, type 2 (T helper 2-mediated) inflammation, and type 3 (T helper 17-mediated) inflammation, which can each be targeted by biologic agents[3]. Analyzing the expression of cytokines, cytokine receptors, and immune checkpoint genes could help us better understand disease-specific patterns of inflammation and their key mediators. In myositis, this could lead to the use of additional more targeted biologic agents, in addition to the current strategies of autoantibody depletion (e.g., rituximab, intravenous immunoglobulin) and targeting of a common cytokine receptor signaling pathway (e.g., Janus kinase inhibitors)[4]. For IBM in particular, where no immunosuppressants or other medications have been shown to be effective, identifying possible immunological targets is of special importance[5].

Thus far, studies of cytokine expression in skeletal muscle of myositis patients have been few and limited in scope[6–13]. A notable exception is the IFN-I pathway, which has been shown in multiple studies to be overactive in the muscle of juvenile and adult DM patients compared to other myositis patients and controls[6, 7, 14–17]. We previously showed that this is mainly driven by *IFNB1*, whose transcript is detectable in muscle of >60% of DM patients and whose protein product (IFN-β) drives the expression of itself and IFN-I-inducible genes in cultured human myoblasts[18]. In addition, *IFNG*, the only type II IFN and an important mediator in type 1 inflammation, as well as IFN-II-inducible genes have been shown to be overexpressed in the muscle of IBM, ASyS, and DM patients as compared to IMNM and control patients[6–11, 16, 17]. Two of these studies examined muscle expression of a large number of cytokines in IBM, identifying cytokines overexpressed in IBM compared to healthy controls and other myositis patients. These included several cytokines involved in type 1 inflammation, such as *CCL5*, *CXCL9*, *CXCL10*, *CXCL11*, and *TNF*[6, 8]. However, there remains a need to better characterize the cytokines and cytokine receptors specific to IBM and other myositis groups.

In this study, we perform a comparison of the expression of 338 genes encoding cytokines, cytokine receptors, and immune checkpoints in the muscle biopsies of 669 myositis patients stratified by clinical group and autoantibody subgroup, in order to better characterize group-specific patterns of inflammation and identify potential therapeutic targets.

## METHODS

### Patients

The classification process of skeletal muscle biopsies from 669 patients into clinical groups and autoantibody subgroups is outlined in the Supplemental Methods, with the classifications listed in Table 1[19–21].

**Table 1.** Patient samples analyzed by clinical group and autoantibody subgroup. Italicized groups were not analyzed separately but were included when calculating the differential expression of genes in groups of interest.

| Clinical Group | Sample Size | Autoantibody Subgroup | Sample Size |
| --- | --- | --- | --- |
| Dermatomyositis (DM) | 105 | Anti-Mi2 (Mi2) | 22 |
|  |  | Anti-MDA5 (MDA5) | 11 |
|  |  | Anti-NXP2 (NXP2) | 21 |
| | | Anti-TIF1γ (TIF1) | 28 |
|  |  | *Other: Anti-SAE, Seronegative* | 23 |
| Immune-mediated necrotizing myopathy (IMNM) | 80 | Anti-HMGCR (HMGCR) | 60 |
|  |  | Anti-SRP (SRP) | 20 |
| Anti-synthetase syndrome (ASyS) | 65 | Anti-Jo1 (Jo1) | 37 |
|  |  | *Other: Anti-PL7, Anti-PL12, Anti-EJ, Anti-OJ* | 28 |
| Inclusion Body Myositis (IBM) | 53 | - |  |
| Anti-PM/Scl Overlap Myositis (PM/Scl) | 19 | Anti-PM/Scl (PM/Scl) | 19 |
| *Other Inflammatory Myopathies* | 239 | - |  |
| *Genetic Myopathies* | 71 | - |  |
| Normal Tissue (NT) | 37 | - |  |
| **Total** | **669** |  |  |

### RNA sequencing

Bulk RNA-seq was performed on frozen muscle biopsy specimens as previously described[2, 16, 22–25]. Briefly, muscle biopsies underwent immediate flash freezing and were stored at -80°C across all contributing centers. Samples were then transported in dry ice to the NIH and processed uniformly to prepare the library and conduct the analysis. RNA was extracted with TRIzol. Libraries were either prepared with the NeoPrep system according to the TruSeq Stranded mRNA Library Prep protocol (Illumina, San Diego, CA) or with the NEBNext Poly(A) mRNA Magnetic Isolation Module and Ultra^™^ II Directional RNA Library Prep Kit for Illumina (New England BioLabs, ref. #E7490, and #E7760).

### Gene Selection

The selection process of 338 genes of interest, encoding cytokines, cytokine receptors, and immune checkpoints is outlined in the Supplementary Methods, with the genes listed and categorized in Supplementary Tables S1-S2[3, 26–31].

### Statistical and bioinformatic analysis

For RNA-seq analysis, sequencing reads were demultiplexed using bcl2fastq/2.20.0 and preprocessed using fastp/0.23.4. The abundance of each gene was determined using Salmon/1.5.2. Counts were normalized using the Trimmed Means of M values (TMM) from edgeR/4.2.1 for graphical analysis. Differential expression was performed using limma/3.60.6. The Benjamin-Hochberg correction was used to adjust for multiple comparisons if appropriate.

The set of genes associated with each myositis group of interest was generated by finding genes that were differentially expressed (*q*-value < 0.001) versus NT and versus all samples, including over- (log-fold change > 0) and under- (log-fold change > 0) expressed genes. The set of genes associated with NT was generated by finding genes that were differentially expressed (*q*-value < 0.001) versus all samples. Venn diagrams were used to represent these analyses graphically. Heatmaps and individual boxplots were used to visualize gene expression for each group. Correlation heatmaps were generated by correlating expression of top overexpressed genes for each group with representative muscle and leukocyte markers, including markers of macrophages, dendritic cells, B cells, and T cell subsets. R and Python software were used to generate all figures.

## RESULTS

### Identification and Quantification of Differentially Expressed Genes by Group

The differentially overexpressed and underexpressed genes for each myositis group and for NT are listed in Table 2. Genes in a given cell of the table are listed from lowest *q*-value vs. all samples to highest (*q*-value < 0.001). In order to visualize the group-specific and group-overlapping genes, Venn Diagram representations of Table 2 were generated (Supplementary Figure S1). Group-specific genes extrapolated from the Venn Diagrams are listed in Table 3.

**Table 2.**
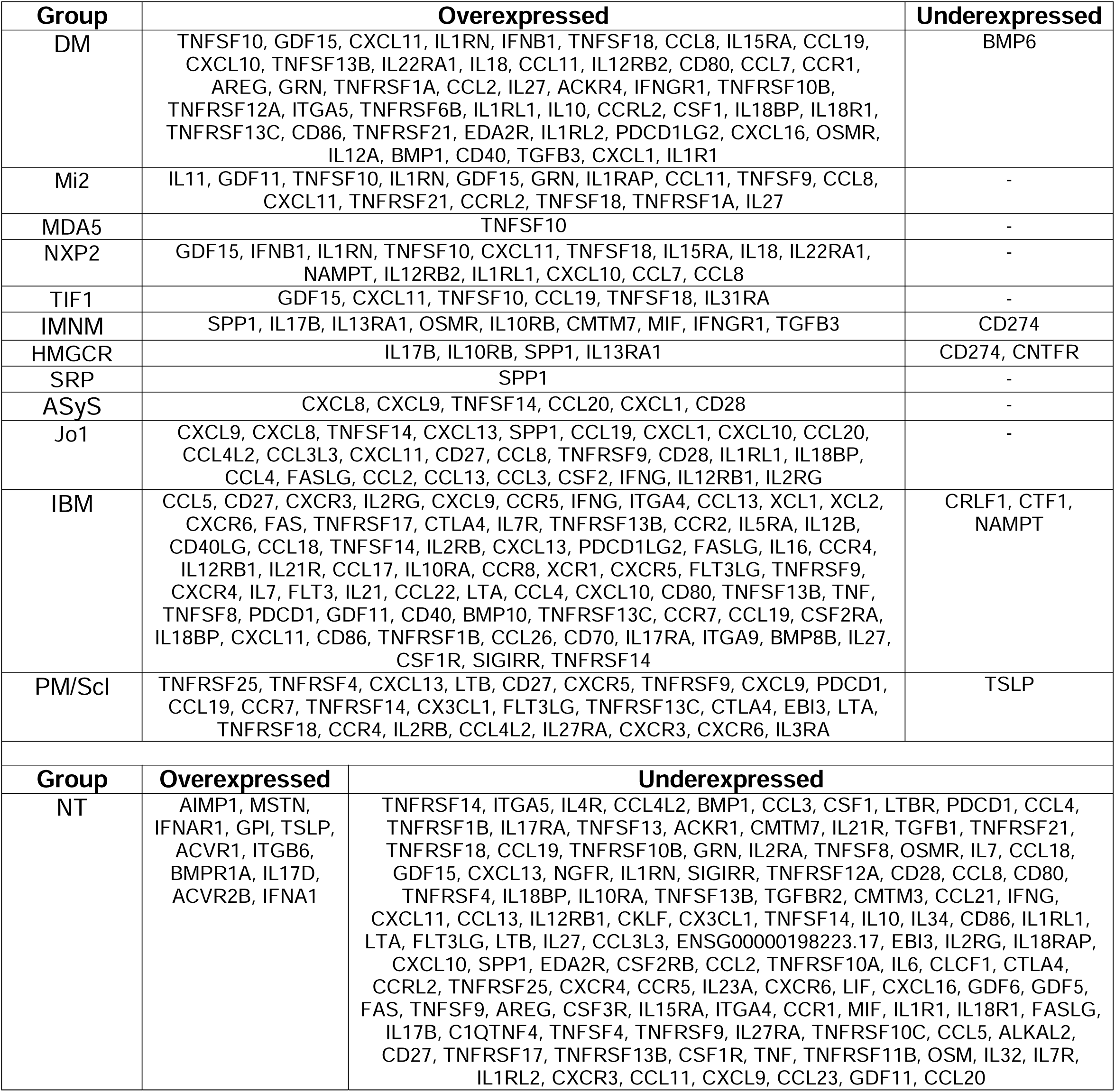
All differentially expressed genes by clinical group and autoantibody subgroup. In order to be included in a given myositis group, genes must have had a *q*-value < 0.001 vs. all samples and vs. NT. In order to be included for NT, genes must have had a *q*-value < 0.001 vs. all samples. Genes are listed in order from lowest *q*-value to highest *q*-value vs. all samples.

**Table 3.** Myositis clinical group- and autoantibody subgroup-specific differentially expressed genes. Genes were included for a given myositis clinical group (DM, IMNM, ASyS, IBM, PM/Scl) if differentially expressed in that group and not differentially expressed in any other myositis group, except its autoantibody subgroups (e.g., HMGCR and SRP for IMNM). Genes were included for a given autoantibody subgroup (Mi2, MDA5, NXP2, TIF1, HMGCR, SRP, Jo1) if differentially expressed in that subgroup and not differentially expressed in any other group, except the clinical group it belongs to (e.g., IMNM for HMGCR).

| Group | Overexpressed | Underexpressed |
| --- | --- | --- |
| DM | TNFSF10, GDF15, IL1RN, IFNB1, TNFSF18, IL15RA, IL22RA1, IL18, CCL11, IL12RB2, CCL7, CCR1, AREG, GRN, TNFRSF1A, ACKR4, TNFRSF10B, TNFRSF12A, ITGA5, TNFRSF6B, IL10, CCRL2, CSF1, IL18R1, TNFRSF21, EDA2R, IL1RL2, CXCL16, IL12A, BMP1, IL1R1 | BMP6 |
| Mi2 | IL11, GRN, IL1RAP, CCL11, TNFSF9, TNFRSF21, CCRL2, TNFRSF1A | - |
| MDA5 | - | - |
| NXP2 | IFNB1, IL15RA, IL18, IL22RA1, NAMPT, IL12RB2, CCL7 | - |
| TIF1 | IL31RA | - |
| IMNM | IL17B, IL13RA1, IL10RB, CMTM7, MIF | CD274 |
| HMGCR | IL17B, IL10RB, IL13RA1 | CD274, CNTFR |
| SRP | - | - |
| AS | CXCL8, CCL20, CD28 | - |
| Jo1 | CXCL8, CCL20, CCL3L3, CD28, CCL3, CSF2 | - |
| IBM | CCL5, CCR5, ITGA4, XCL1, XCL2, FAS, TNFRSF17, IL7R, CCR2, IL5RA, IL12B, CD40LG, CCL18, IL16, IL21R, CCL17, IL10RA, CCR8, XCR1, CXCR4, IL7, FLT3, IL21, CCL22, TNFSF13B, TNF, TNFSF8, BMP10, CSF2RA, TNFRSF1B, CCL26, CD70, IL17RA, ITGA9, BMP8B, CSF1R, SIGIRR | CRLF1, CTF1, NAMPT |
| PM/ScI | TNFRSF25, TNFRSF4, LTB, CX3CL1, EBI3, TNFRSF18, IL27RA, IL3RA | TSLP |

Among myositis clinical groups, IBM had the most differentially overexpressed genes (71), followed by DM (49), PM/Scl (26), IMNM (9), and ASyS (6). The autoantibody subgroup with the most differentially overexpressed genes (after PM/Scl) was Jo1 with 27, including all 6 genes differentially overexpressed in the clinical group it belongs to, ASyS. Among the DM autoantibody subgroups, Mi2 had the most differentially overexpressed genes (16), followed by NXP2 (15), TIF1 (6), and MDA5 (1). Among the IMNM autoantibody subgroups, HMGCR had the most differentially overexpressed genes (4), followed by SRP (1) (Table 2).

IBM had the most group-specific differentially overexpressed genes with 37, followed by DM and its autoantibody subgroups (36), PM/Scl (8), ASyS and its autoantibody subgroup (6), and IMNM and its autoantibody subgroups (5) (Supplementary Figure S1A). Among the 36 DM-specific genes, 8 were Mi2-specific, 7 were NXP2-specific, 1 was TIF1-specific, and none were MDA5-specific (Supplementary Figure S1B). Among the 5 IMNM-specific genes, 3 were HMGCR-specific and none were SRP-specific. The largest number of genes differentially overexpressed in more than one group was 11 for the pair IBM-PM/Scl, followed by 7 for the pairs DM-IBM and ASyS-IBM. The largest number of genes differentially overexpressed in three groups was 4 for the trio ASyS-IBM-PM/Scl. As expected by the nature of the analysis, there were no genes differentially overexpressed in all 5 groups. However, there were 117 genes differentially underexpressed in NT vs. all samples, an imperfect surrogate for pan-myositis overexpressed genes. Differentially underexpressed genes for myositis groups were very few, ranging from 0 to 3 per group.

### Genes differentially expressed in DM and correlation with muscle and leukocyte markers

DM patients exhibited differential overexpression of 49 of the 338 genes examined (14%), of which 31 were specific to the DM clinical group (not including genes only overexpressed in a DM autoantibody subgroup but not in the clinical group). The top 5 DM-specific genes were *TNFSF10*, *GDF15* (a member of the TGFB family), *IL1RN* (interleukin-1 receptor antagonist, an IFN-I-inducible gene), *IFNB1*, and *TNFSF18*. *CXCL11*, an IFN-II-inducible chemokine, was the third highest differentially overexpressed gene in DM, but was shared with Jo1 and IBM. The top 15 overexpressed genes in DM were negatively correlated with mature muscle markers (*ACTA1*, *MYH1*, *MYH2*) and mitochondrial genes (*MT-CO1*, *MT-CO2*) and positively correlated with markers of muscle regeneration (*NCAM1*, *MYOG*, *PAX7*, *MYH3*, *MYH8*), indicating correlation with disease activity. The top 15 genes were also generally correlated with T cell markers (*CD3E*, *CD4*, *CD8A*), macrophage markers (*CD68*, *CD14*), and markers of type 1 inflammation (*TBX21*, *STAT1*), more so than markers of type 2 inflammation (*GATA3*, *STAT6*) (Figure 1A). There was only 1 differentially underexpressed gene for DM patients, *BMP6* (another member of the TGFB family).

**Figure 1.**
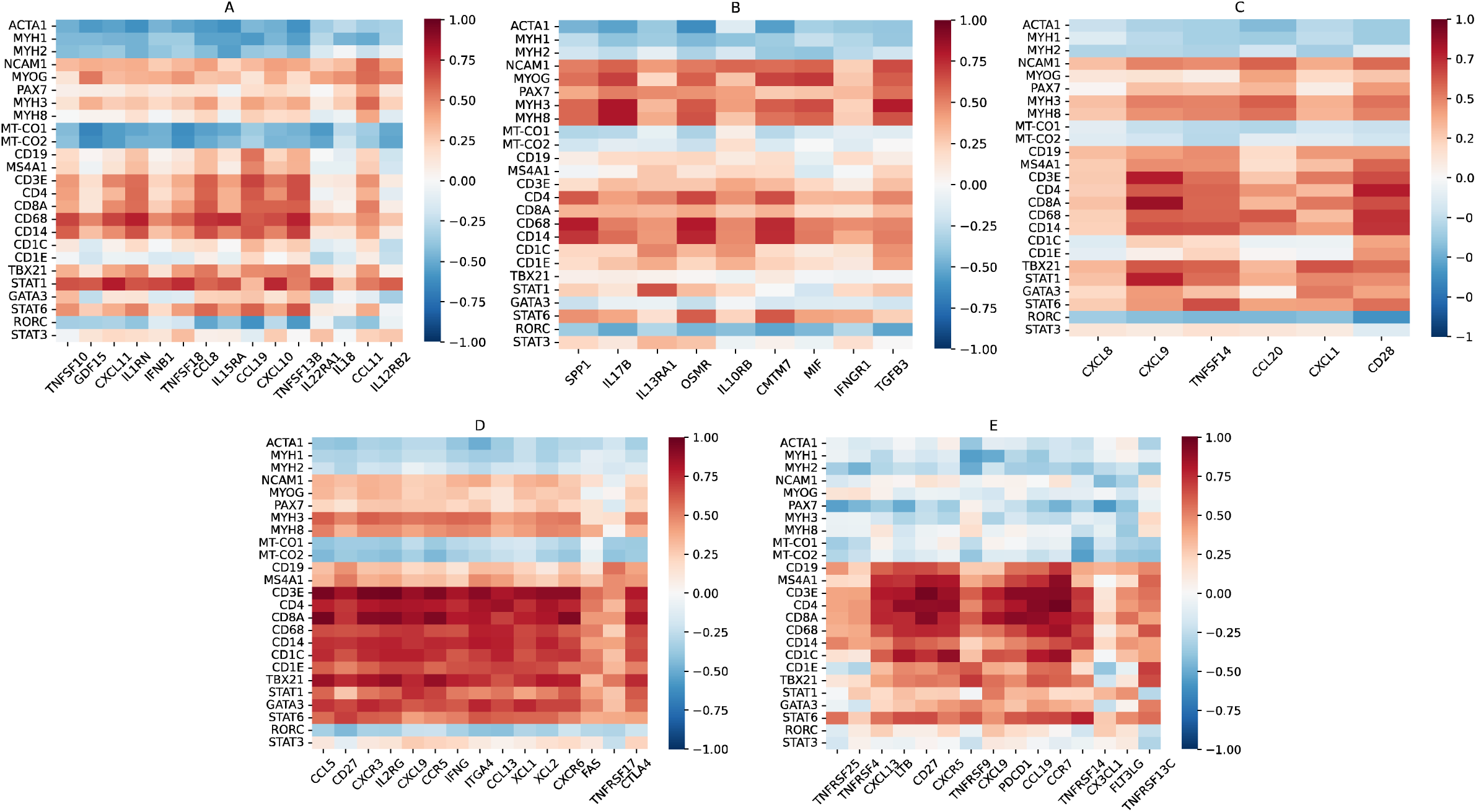
Correlation heatmaps for top differentially overexpressed genes by clinical group vs. muscle and leukocyte markers. For all heatmaps, the x-axis displays the top 15 differentially overexpressed genes for that clinical group, or all differentially overexpressed genes if there are less than 15, ordered from lowest *q*-value vs. all samples to highest. The y-axis displays mature muscle markers (*ACTA1*, *MYH1*, *MYH2*), markers of muscle regeneration (*NCAM1*, *MYOG*, *PAX7*, *MYH3*, *MYH8*), mitochondrial markers (*MT-CO1*, *MT-CO2*), B cell markers (*CD19*, *MS4A1*), T cell markers (*CD3E*, *CD4*, *CD8A*), macrophage markers (*CD14*, *CD68*), dendritic cell markers (*CD1C*, *CD1E*), type 1 markers (*TBX21*, *STAT1*), type 2 markers (*GATA3*, *STAT6*), and type 3 markers (*RORC*, *STAT3*). Heatmaps displayed are for DM (A), IMNM (B), ASyS (C), IBM (D), and PM/Scl (E).

Mi2 patients exhibited differential overexpression of 16 genes, of which 8 were not shared with any other DM autoantibody subgroup or non-DM group. These included 3 genes (*IL11*, *IL1RAP*, and *TNFSF9*) that were differentially overexpressed in Mi2 patients, but not in the DM clinical group, indicating strong Mi2 specificity. We previously established *IL11* as one of the genes that is derepressed following the internalization of anti-Mi2 autoantibodies[25]. The top 15 differentially overexpressed genes in Mi2 patients exhibited characteristically strong negative correlation with mature muscle and mitochondrial markers and strong positive correlation with markers of muscle regeneration. They were also positively correlated with leukocyte markers, including B cell markers (*CD19*, *MS4A1*), T cell markers, macrophage markers, and markers of type 1 inflammation more so than markers of type 2 inflammation (Figure 2A).

**Figure 2.**
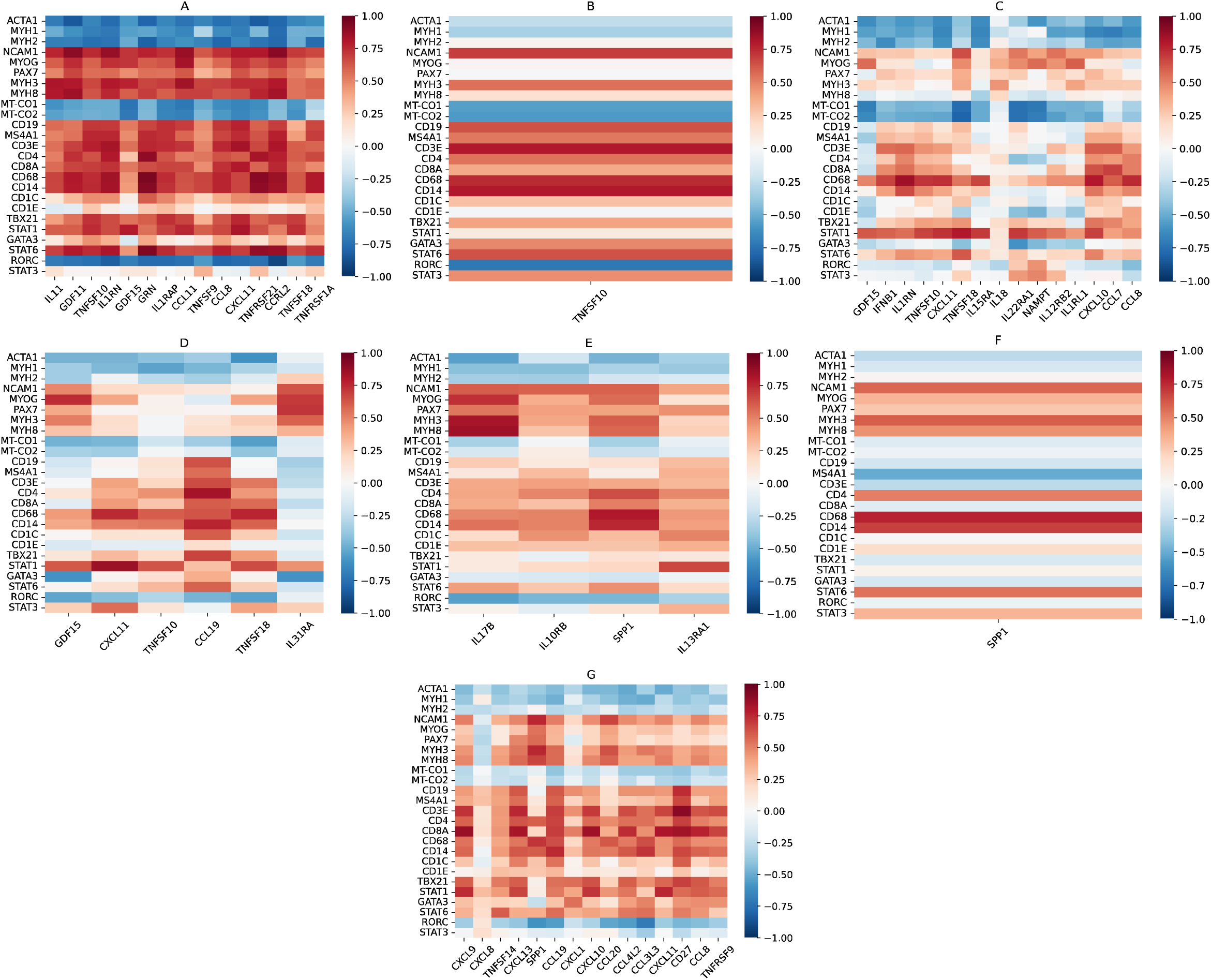
Correlation heatmaps for top differentially overexpressed genes by autoantibody subgroup vs. muscle and leukocyte markers. For all heatmaps, the x-axis displays the top 15 differentially overexpressed genes for that autoantibody subgroup, or all differentially overexpressed genes if there are less than 15, ordered from lowest *q*-value vs. all samples to highest. The y-axis displays mature muscle markers (*ACTA1*, *MYH1*, *MYH2*), markers of muscle regeneration (*NCAM1*, *MYOG*, *PAX7*, *MYH3*, *MYH8*), mitochondrial markers (*MT-CO1*, *MT-CO2*), B cell markers (*CD19*, *MS4A1*), T cell markers (*CD3E*, *CD4*, *CD8A*), macrophage markers (*CD14*, *CD68*), dendritic cell markers (*CD1C*, *CD1E*), type 1 markers (*TBX21*, *STAT1*), type 2 markers (*GATA3*, *STAT6*), and type 3 markers (*RORC*, *STAT3*). Heatmaps displayed are for Mi2 (A), MDA5 (B), NXP2 (C), TIF1 (D), HMGCR (E), SRP (F), and Jo1 (G).

MDA5 patients only exhibited differential overexpression of 1 gene (*TNFSF10*), but this was not MDA5-specific. *TNFSF10* was inversely correlated with mature muscle markers and mitochondrial markers, and strongly positively correlated with *CD3E* and macrophage markers in MDA5 patients (Figure 2B).

NXP2 patients exhibited differential overexpression of 15 genes, of which 7 were not shared with any other DM autoantibody subgroup or non-DM group. These included 1 gene (*NAMPT*) that was differentially overexpressed in NXP2 patients, but not in the DM clinical group, indicating strong NXP2 specificity. The 15 differentially overexpressed genes in NXP2 also exhibited correlation with muscle markers and leukocyte markers, including macrophage markers and markers of type 1 inflammation, but correlations were weaker than for Mi2 patients (Figure 2C).

TIF1 patients exhibited differential overexpression of 6 genes, of which 1 was not shared with any other group (*IL31RA*). The top differentially overexpressed genes in TIF1 exhibited similar levels of correlation with muscle markers and leukocyte markers as for NXP2 patients (Figure 2D).

### Genes differentially overexpressed in IMNM and correlation with muscle and leukocyte markers

The IMNM patient group exhibited differential overexpression of only 9 of the 338 genes examined (3%). The top gene was *SPP1* (osteopontin), which was also overexpressed in Jo1 patients. There were 5 IMNM-specific genes, including *IL17B*, *IL13RA1*, *IL10RB*, *CMTM7*, and *MIF*. All 9 genes were negatively correlated with mature muscle markers and mitochondrial genes and positively correlated with markers of muscle regeneration, indicating correlation with disease activity. There was less correlation with leukocyte markers, but *SPP1*, *OSMR*, and *CMTM7* were notably well correlated with *CD4* and macrophage markers. There was no strong correlation between the 9 genes and markers of T subsets, indicating a lack of predominant type 1, type 2, or type 3 inflammation (Figure 1B). There was one gene differentially underexpressed in IMNM patients, the immune checkpoint *CD274* (PD-L1).

HMGCR patients exhibited differential overexpression of 4 genes (*IL17B*, *IL10RB*, *SPP1*, *IL13RA1*), which were all also differentially overexpressed in the IMNM clinical group. As with IMNM, these genes were correlated well with muscle markers, while *SPP1* was strongly correlated with *CD4* and macrophage markers (Figure 2E). SRP patients exhibited differential overexpression of only 1 gene (*SPP1*). The same correlations between *SPP1* and muscle and leukocyte markers were seen in the SRP group (Figure 2F).

### Genes differentially overexpressed in ASyS and correlation with muscle and leukocyte markers

As mentioned previously, there were 27 genes differentially overexpressed in Jo1 patients, which included all 6 genes differentially overexpressed in ASyS patients, suggesting that Jo1 was the main source of this transcriptomic pattern. Of the 27 genes, there were only 6 that were ASyS/Jo1-specific (*CXCL8*, *CCL20*, *CCL3L3*, *CD28*, *CCL3*, and *CSF2*), with many overexpressed genes shared with the IBM (15), DM (8), and PM/Scl (6) groups. This included the top differentially overexpressed gene in Jo1 (*CXCL9*), which was shared with IBM and PM/Scl. The top 15 differentially overexpressed genes in Jo1 were negatively correlated with mature muscle markers and mitochondrial markers, while all but *CXCL8* were positively correlated with markers of muscle regeneration. The same genes were also generally correlated with leukocyte markers, especially for *CD8* and markers of type 1 inflammation more than type 2 inflammation (Figure 2G). The same correlations were seen in the ASyS clinical group for the 6 overexpressed genes in ASyS patients (Figure 1C).

### Genes differentially overexpressed in IBM and correlation with muscle and leukocyte markers

IBM patients exhibited differential overexpression of 71 of the 338 genes examined (21%), of which 37 were specific to IBM. The top 5 IBM-specific genes were *CCL5*, *CCR5* (the receptor for CCL5), *ITGA4*, *XCL1*, and *XCL2*. *XCR1*, a dendritic cell marker and the receptor for XCL1 and XCL2, was also specifically differentially overexpressed in IBM. The top 7 differentially overexpressed genes in IBM also included *CD27* (a co-stimulatory protein on T cells), *CXCR3* (a T_H_1/T_C_1 marker), *IL2RG* (a common cytokine receptor subunit), the IFN-II-inducible chemokine *CXCL9*, and *IFNG*. The top 15 differentially overexpressed genes in IBM were negatively correlated with mature muscle markers and mitochondrial markers, while all but *TNFRSF17* were positively correlated with markers of muscle regeneration. The same genes were also strongly correlated with leukocyte markers, especially *CD8* and type 1 inflammatory cells slightly more than type 2 inflammatory cells, but also macrophages and dendritic cells (Figure 1D). There were three differentially underexpressed genes in IBM patients, *CRLF1*, *CTF1*, and *NAMPT*.

### Genes differentially overexpressed in PM/Scl and correlation with muscle and leukocyte markers

PM/Scl patients exhibited differential overexpression of 26 of the 338 genes examined (8%), of which 8 were specific to PM/Scl. The top 5 PM/Scl-specific genes were *TNFRSF25*, *TNFRSF4*, *LTB*, *CX3CL1*, and *EBI3* (a common subunit of IL-27 and IL-35). The top 5 differentially overexpressed genes in PM/Scl also included *CXCL13* and *CD27*. The top 15 differentially overexpressed genes in PM/Scl were generally negatively correlated with mature muscle markers and mitochondrial markers, but also largely negatively correlated or not correlated with markers of muscle regeneration. The same genes were strongly correlated with leukocytes including T cells (both type 1 and type 2 inflammation), macrophages, and dendritic cells (Figure 1E). There was one differentially underexpressed gene in PM/Scl, the type 2 alarmin cytokine *TSLP*.

### Genes associated with type 1 inflammation are comparatively enriched in IBM, PM/Scl, Jo1, and DM patients

Given the observed correlations of overexpressed genes and markers of type 1 and type 2 inflammation, we also sought to compare the expression of markers of type 1, type 2, and type 3 inflammation we previously defined in Supplementary Table S2. For each gene, the *q*-value of differential overexpression and log-fold change for each group are shown in Supplementary Tables S3-S5. For visualization, we generated a heatmap of the median expression of these genes for each of the autoantibody subgroups, IBM, and NT (Figure 3).

**Figure 3.**
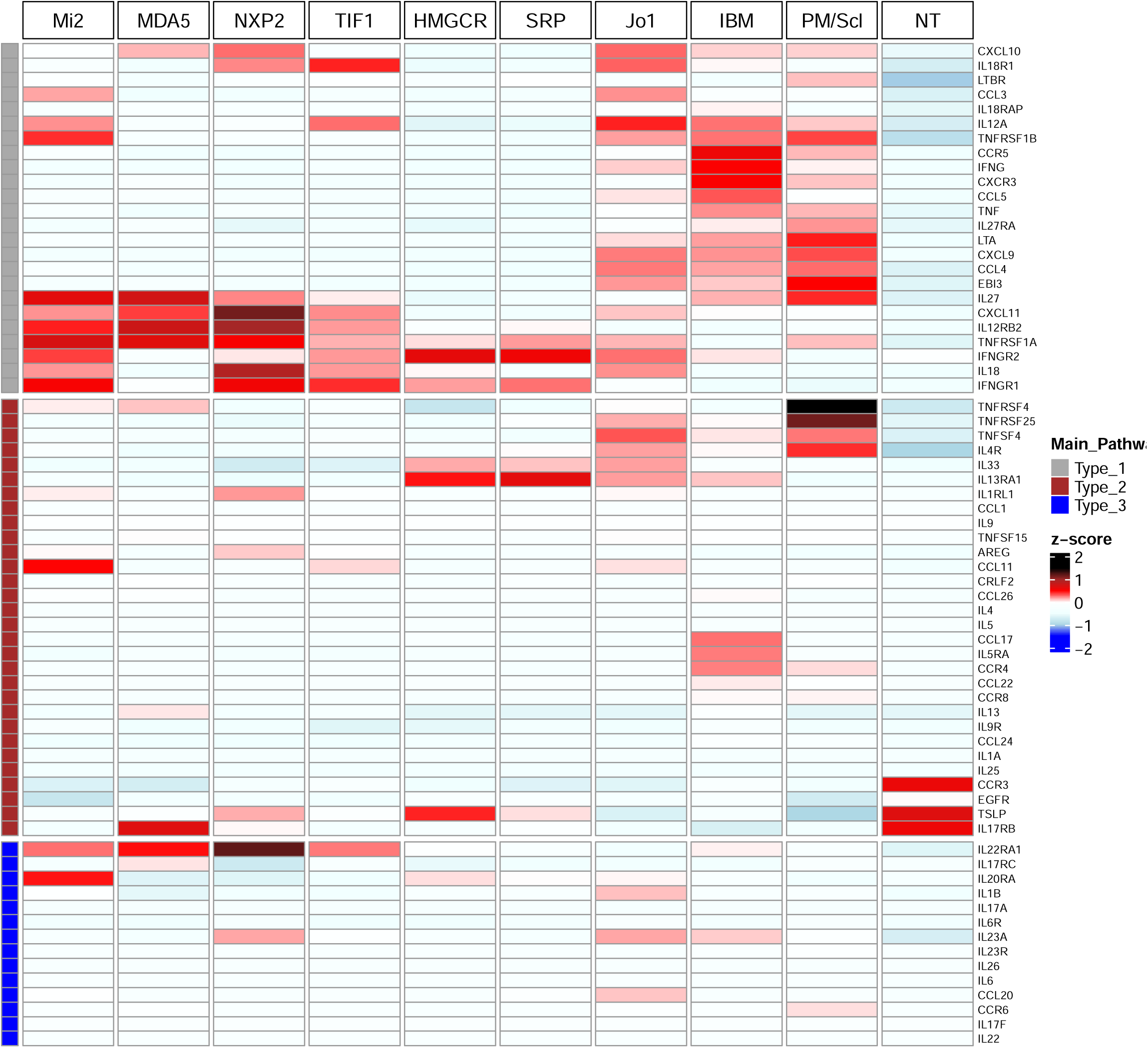
Heatmap of type 1, type 2, and type 3 gene expression by group. The heatmap displays median expression levels of type 1, type 2, and type 3 genes for each group. Groups included are all autoantibody subgroups (Mi2, MDA5, NXP2, TIF1, HMGCR, SRP, Jo1, PM/Scl), as well as IBM (no autoantibody subgroup) and NT (control).

Expression of markers associated with type 1 inflammation predominated over markers of type 2 and type 3 inflammation in DM, Jo1, IBM, and PM/Scl patients, while none of the three were particularly overexpressed in IMNM (Figure 3). In particular, Jo1, IBM, and PM/Scl patients exhibited shared overexpression of a particular set of markers associated with type 1 inflammation, generally distinct from the set overexpressed in DM patients (Figure 3, Supplemental Table 3). IBM patients had the most differentially overexpressed markers of type 1 inflammation (12), of which 2/3 were also differentially overexpressed in one or more other groups (*CCL4*, *CXCL9*, *CXCL10*, *CXCL11*, *CXCR3*, *IFNG*, *IL27*, and *LTA*). The most IBM-specific markers of type 1 inflammation were the ligand-receptor pair *CCL5*-*CCR5* (first and sixth most differentially overexpressed genes in IBM), while IBM patients also exhibited markedly higher median expression of the T_H_1/T_C_1 marker *CXCR3* and *IFNG* than other groups. PM/Scl patients exhibited markedly higher median expression of *LTA* and *EBI3* than other groups. Notably, there were four differentially overexpressed markers of type 1 inflammation unique to DM patients (*IL12RB2*, *TNFRSF1A*, and the ligand-receptor pair *IL18*-*IL18R1*), while they also exhibited markedly higher median expression of *CXCL11* than the other groups.

Markers of type 2 inflammation were generally less commonly overexpressed among myositis groups (Figure 3, Supplemental Table 4). A notable exception was PM/Scl, where the top two differentially overexpressed genes, *TNFRSF25* and *TNFRSF4*, are both implicated in type 2 inflammation. Other notable markers of type 2 inflammation that were group-specific were *AREG* (DM), *CCL11* (DM), *IL13RA1* (IMNM), *CCL17* (IBM), and *IL5RA* (IBM). Markers of type 3 inflammation were even less commonly overexpressed among the myositis groups, with exceptions being *IL22RA1* in DM and *CCL20* in Jo1 (Figure 3, Supplemental Table 5).

## DISCUSSION

Our findings add support to previous work implicating mediators of type 1 inflammation in the pathogenesis of IBM. For example, it has been demonstrated that highly differentiated *CD8*+ T cells are elevated in muscle and peripheral blood of IBM patients, including *TBX21*+ *CD8*+ T cells (T_C_1 cells) in particular[8, 11, 32, 33]. Previous studies have also shown elevated muscle levels of numerous cytokines associated with type 1 inflammation, including *CCL5*, *CXCL9*, *CXCL10*, *CXCL11*, *IFNG*, and *TNF* in IBM patients compared to various myositis and non-myositis controls[6, 8–11]. Similar elevations of cytokines associated with type 1 inflammation, including CXCL9, CXCL10, IFN-γ, IL-12, and TNF have been demonstrated at the protein level in serum of IBM patients versus non-myositis controls[11, 34]. In our study, we confirmed the elevation of these cytokines in IBM, while also finding a strong correlation between the most differentially overexpressed inflammatory mediators in IBM and *CD8A* and *TBX21*. Furthermore, we found that the T_H_1/T_C_1 markers *CXCR3* and *CCR5* were differentially overexpressed in IBM, with higher median expression than in any other myositis group. The *CCL5*-*CCR5* ligand-receptor pair was particularly specific for IBM, representing the top two IBM-specific differentially overexpressed genes. Interestingly, an allelic variant of *CCR5* resulting in a non-functional receptor (CCR5Δ32) is thought to be protective against IBM according to a large genetic association study[35]. Furthermore, CCR5 is a macrophage entry receptor for R5-tropic strains of the human immunodeficiency virus (HIV), which has been associated with IBM[36]. Taken together, the *CCL5*-*CCR5* axis seems to be particularly important to the type 1 inflammation in IBM and could represent a therapeutic target. CCR5 inhibitors are indicated for HIV infection and have been in clinical trials for a variety of other autoimmune diseases, including rheumatoid arthritis, graft-versus-host disease, and primary sclerosing cholangitis[37]. Furthermore, antigen-presenting dendritic cells have also been implicated in IBM inflammation[38]. Here, we found the type 1 conventional dendritic cell marker *XCR1* and its ligands, *XCL1* and *XCL2*, to be overexpressed in IBM patients and IBM-specific. The *XCL1*-*XCR1* interaction activates T_C_1 cells and has been implicated in other autoimmune disorders with type 1 inflammation, including sarcoidosis, Crohn’s disease, and rheumatoid arthritis [39, 40]. Although IBM has proved to be refractory to corticosteroids and some other immunosuppressants, this may be due to the durable nature of highly differentiated *CD8*+ T cells in evading cell death mechanisms, and they may be more susceptible to more targeted therapeutics[5].

Our findings also demonstrated a similar overexpression of markers of type 1 inflammation in Jo1 and PM/Scl patients, albeit weaker than in IBM. For Jo1, the IFN-II-inducible chemokine *CXCL9* was the top differentially overexpressed gene, while the neutrophil chemoattractant *CXCL8* was the most Jo1-specific differentially overexpressed gene. Both have been previously shown to be elevated in serum of ASyS patients compared to healthy controls[41, 42]. In PM/Scl, besides type 1 genes, we identified several PM/Scl-specific differentially overexpressed genes, including *TNFRSF25* and *TNFRSF4*. In DM, our data also revealed a strong type 1 inflammation pattern, but the specific type 1 genes that were most differentially overexpressed (e.g., *IL18* and *IL12RB2*) were different from those in IBM, Jo1, and PM/Scl. *IFNB1* was the fourth most differentially overexpressed gene analyzed in DM, consistent with our previous results demonstrating a strong type I IFN signature in DM[16, 18]. The top three DM-specific differentially overexpressed genes were *TNFSF10*, *GDF15*, and *IL1RN*, which have all been studied as potential biomarkers for DM[43–45]. Finally, IMNM had the least number of differentially overexpressed genes among myositis clinical groups with no predominant inflammatory pattern.

Our study has several limitations. Firstly, we only measured RNA, so the results reflect muscle expression of cytokines rather than circulating cytokines. Secondly, we used bulk RNA sequencing, so we were not able to distinguish cellular sources of gene expression. Thirdly, sample sizes between groups varied, which impacted *q*-values. The variable sample sizes also meant that some groups were weighed more than others when calculating differential expression. Finally, the differential expression analysis was designed to identify group-specific genes, rather than common overexpressed genes among multiple groups.

In conclusion, we identified differentially overexpressed cytokines, cytokine receptors, and immune checkpoints in different myositis clinical groups and autoantibody subgroups. Our results point to a predominance of type 1 inflammation in IBM, Jo1, and PM/Scl, while DM patients exhibited overexpression of a different set of type 1 inflammatory genes, in addition to a strong Type I IFN signature. Our results also highlight the *CCL5*-*CCR5* and *XCL1*-*XCL2*-*XCR1* axes as especially IBM-specific, potentially representing therapeutic targets.

## Acknowledgments

Julie Thompson for her invaluable help maintaining the NIH Natural History Protocol, the NIAMS Sequencing Core, and its members.

## Contributorship

All authors contributed to the development of the manuscript, including interpretation of results, substantive review of drafts, and approval of the final draft for submission.

## Ethical approval information

All biopsies were from subjects enrolled in institutional review board (IRB)-approved longitudinal cohorts in the National Institutes of Health, the Johns Hopkins, the Clinic Hospital, the Vall d’Hebron Hospital, the Mayo Clinic, and the Charité-Universitätsmedizin Berlin.

## Patient and public involvement

Patients and/or the public were not involved in the design, conduct, reporting, or dissemination plans of this research.

## Data availability statement

Any anonymized data not published within the article will be shared by request from any qualified investigator.

