## Supplementary Material for "Distinct Cytokine and Cytokine Receptor Expression Patterns Characterize Different Forms of Myositis"

**Supplementary Methods.**

### Patients

Muscle tissue from 669 patients who underwent diagnostic muscle biopsies and healthy volunteers at several centers specialized in neuromuscular diseases underwent bulk RNA sequencing (RNA-seq) analysis. Muscle biopsies from myopathy patients were obtained from institutional review board-approved longitudinal cohorts from the National Institutes of Health in Bethesda, MD, the Johns Hopkins Myositis Center in Baltimore, MD, the Vall d'Hebron and Clinic Hospitals in Barcelona, Spain, the Mayo Clinic in Rochester, MN, and the Charité-Universitätsmedizin in Berlin, Germany.

All samples were divided into clinical groups, including DM, IMNM, ASyS, IBM, overlap myositis, other inflammatory myopathies, genetic myopathies, and normal tissue (NT). Some samples were also classified into autoantibody subgroups, including Mi2, MDA5, NXP2, TIF1, HMGCR, SRP, Jo1, and PM/Scl (Table 1). Since the only overlap myositis samples analyzed were from anti-PM/Scl patients, we will refer to PM/Scl as both a clinical group and an autoantibody subgroup, in order to compare it to other myopathies from both categories. Patients were classified as IBM if they fulfilled Lloyd's criteria for inclusion body myositis[1]. Patients testing positive for a myositis-specific autoantibody were classified according to the Casal and Pinal criteria[2]. The same criteria were used to classify patients as PM/Scl, although anti-PM/Scl autoantibodies are myositis-associated autoantibodies. Patients who met the 2017 ACR/EULAR classification criteria for DM, but were negative for DM-specific autoantibodies were also classified as DM[3].

Autoantibody testing was performed using one or more of the following techniques: ELISA, immunoprecipitation of proteins generated by *in vitro* transcription and translation (IVTT-IP), line blotting (EUROLINE myositis profile), or immunoprecipitation from 35S-methionine-labeled HeLa cell lysates.

### Standard protocol approvals and patient consent

This study was approved by the Institutional Review Boards of the National Institutes of Health, the Johns Hopkins University, the Vall d'Hebron University Hospital, the Mayo Clinic, and the Charité-Universitätsmedizin Berlin. Written informed consent was obtained from each participant. All methods were performed in accordance with the relevant guidelines and regulations.

### Gene Selection

A total of 338 genes of interest were chosen, including 200 cytokines, 131 cytokine receptors, and 7 immune checkpoints (Supplementary Table S1). Cytokines included all protein-coding genes in a set of HUGO Gene Nomenclature groups (chemokine ligands, interleukins, interleukin-6 family, interferons, and tumor necrosis factor superfamily), all genes with the UniProt molecular function keyword "cytokine", and four additional cytokines that did not meet these criteria (*CRLF1*, *IL22RA2*, *KITLG*, and *TXLNA*). Two cytokines identified from HUGO (*CCL3L1* and *CCL4L2*) that were not present in our reference genome (GRCh38) were excluded. Cytokine receptors included all protein-coding genes in a set of HUGO Gene Nomenclature groups (chemokine receptors, interleukin receptors, interferon receptors, and tumor necrosis factor receptor superfamily), gene matches for the selected cytokines found in both of two receptor-ligand databases (Ramilowski 2015 & CellphoneDB v5.0.0), and three additional receptors that did not meet these criteria (*CRLF2*, *IL18BP*, *SIGIRR*)[4, 5]. Immune checkpoint genes included were *CD28*, *CD80*, *CD86*, *CD274*, *CTLA4*, *PDCD1*, and *PDCD1LG2*.

All cytokines and cytokine receptors were classified into families, including chemokines, interferons, interleukins, tumor necrosis factor (TNF), transforming growth factor-β (TGFB), and others (Supplementary Table S1). A subset of genes was also classified by the type of inflammation in which they are predominantly overexpressed, as defined by subsets of T cells, innate lymphoid cells (ILCs), and other interacting cells (Supplementary Table S2)[6-10]. These classifications were type 1 (mediated by *TBX21*+ ILC1 cells, $T_H1$ cells, and $T_C1$ cells), type 2 (mediated by *GATA3*+ ILC2 cells, $T_H2$ cells, and $T_C2$ cells), and type 3 (mediated by *RORC*+ ILC3 cells, $T_H17$ cells, and $T_C17$ cells)[6].

**Supplementary Table S1. List of genes included (n=338) by family.**

| Family | Cytokines | Receptors |
|---|---|---|
| Chemokines (and related) | CCL1, CCL2, CCL3, CCL3L3, CCL4, CCL4L2, CCL5, CCL7, CCL8, CCL11, CCL13, CCL14, CCL15, CCL16, CCL17, CCL18, CCL19, CCL20, CCL21, CCL22, CCL23, CCL24, CCL25, CCL26, CCL27, CCL28, CKLF, CMTM1, CMTM2, CMTM3, CMTM5, CMTM7, CMTM8, CX3CL1, CXCL1, CXCL2, CXCL3, CXCL5, CXCL6, CXCL8, CXCL9, CXCL10, CXCL11, CXCL12, CXCL13, CXCL14, CXCL16, CXCL17, PF4, PF4V1, PPBP, TAFA5, XCL1, XCL2 | ACKR1, ACKR2, ACKR3, ACKR4, CCR1, CCR2, CCR3, CCR4, CCR5, CCR6, CCR7, CCR8, CCR9, CCR10, CCRL2, CX3CR1, CXCR1, CXCR2, CXCR3, CXCR4, CXCR5, CXCR6, HRH4, PITPNM3, XCR1 |
| Interferons | IFNA1, IFNA2, IFNA4, IFNA5, IFNA6, IFNA7, IFNA8, IFNA10, IFNA13, IFNA14, IFNA16, IFNA17, IFNA21, IFNB1, IFNE, IFNG, IFNK, IFNL1, IFNL2, IFNL3, IFNW1 | IFNAR1, IFNAR2, IFNGR1, IFNGR2, IFNLR1 |
| Interleukins (and associated) | CLCF1, CNTF, CRLF1, CSF2, CTF1, C17ORF99, EBI3, IL1A, IL1B, IL1F10, IL1RN, IL2, IL3, IL4, IL5, IL6, IL7, IL9, IL10, IL11, IL12A, IL12B, IL13, IL15, IL16, IL17A, IL17B, IL17C, IL17D, IL17F, IL18, IL19, IL20, IL21, IL22, IL22RA2, IL23A, IL24, IL25, IL26, IL27, IL31, IL32, IL33, IL34, IL36A, IL36B, IL36G, IL36RN, IL37, LIF, OSM, TSLP, TXLNA | CNTFR, CRLF2, CSF2RA, CSF2RB, IL1R1, IL1R2, IL1RAP, IL1RAPL1, IL1RAPL2, IL1RL1, IL1RL2, IL2RA, IL2RB, IL2RG, IL3RA, IL4R, IL5RA, IL6R, IL6ST, IL7R, IL9R, IL10RA, IL10RB, IL11RA, IL12RB1, IL12RB2, IL13RA1, IL13RA2, IL15RA, IL17RA, IL17RB, IL17RC, IL17RD, IL17RE, IL18BP, IL18R1, IL18RAP, IL20RA, IL20RB, IL21R, IL22RA1, IL23R, IL27RA, IL31RA, LIFR, OSMR, SIGIRR |
| TNF | C1QTNF4, CD40LG, CD70, EDA, FASLG, LTA, LTB, TNF, TNFSF4, TNFSF8, TNFSF9, TNFSF10, TNFSF11, TNFSF12, TNFSF13, TNFSF13B, TNFSF14, TNFSF15, TNFSF18 | CD27, CD40, EDA2R, EDAR, FAS, LTBR, NGFR, RELT, TNFRSF1A, TNFRSF1B, TNFRSF4, TNFRSF6B, TNFRSF8, TNFRSF9, TNFRSF10A, TNFRSF10B, TNFRSF10C, TNFRSF10D, TNFRSF11A, TNFRSF11B, TNFRSF12A, TNFRSF13B, TNFRSF13C, TNFRSF14, TNFRSF17, TNFRSF18, TNFRSF19, TNFRSF21, TNFRSF25 |
| TGFB | BMP1, BMP2, BMP3, BMP4, BMP5, BMP6, BMP7, BMP8A, BMP8B, BMP10, BMP15, CER1, GDF1, GDF2, GDF3, GDF5, GDF6, GDF7, GDF9, GDF10, GDF11, GDF15, LEFTY1, LEFTY2, MSTN, NODAL, TGFB1, TGFB2, TGFB3 | ACVR1, ACVR1B, ACVR1C, ACVR2A, ACVR2B, ACVRL1, BMPR1A, BMPR1B, BMPR2, ITGAV, ITGB6, ITGB8, TGFBR1, TGFBR2, TGFBR3 |
| Other | AIMP1, ALKAL1, ALKAL2, AREG, CSF1, CSF3, FAM3B, FLT3LG, GPI, GPR15LG, GREM1, GREM2, GRN, KITLG, MIF, MSMP, NAMPT, SCGB3A1, SLURP1, SPP1, THNSL2, THPO, VSTM1 | CSF1R, CSF3R, EGFR, FLT3, ITGA4, ITGA5, ITGA9, ITGB1, KIT, MPL |
| Immune Checkpoint | CD28, CD80, CD86, CD274, CTLA4, PDCD1, PDCD1LG2 | |

**Supplementary Table S2. List of genes classified as type 1, type 2, or type 3.** Only including genes predominantly associated with one of the three inflammation types. Excluding genes commonly involved in multiple inflammation types (e.g., IL2).

| Inflammation Type | Cytokines | Receptors |
|---|---|---|
| ILC1/$T_H$1/$T_C$1-associated (Type 1) | CCL3, CCL4, CCL5, CXCL9, CXCL10, CXCL11, EBI3, IFNG, IL12A, IL18, IL27, LTA, TNF | CCR5, CXCR3, IFNGR1, IFNGR2, IL12RB2, IL18R1, IL18RAP, IL27RA, LTBR, TNFRSF1A, TNFRSF1B |
| ILC2/$T_H$2/$T_C$2-associated (Type 2) | AREG, CCL1, CCL11, CCL17, CCL22, CCL24, CCL26, IL1A, IL4, IL5, IL9, IL13, IL25, IL33, TNFSF4, TNFSF15, TSLP | CCR3, CCR4, CCR8, CRLF2, EGFR, IL1RL1, IL4R, IL5RA, IL9R, IL13RA1, IL17RB, TNFRSF4, TNFRSF25 |
| ILC3/$T_H$17/$T_C$17-associated (Type 3) | CCL20, IL1B, IL6, IL17A, IL17F, IL22, IL23A, IL26 | CCR6, IL6R, IL17RC, IL20RA, IL22RA1, IL23R |

**Supplementary Table S3. Adjusted p-values (q-values) of differential overexpression and log-fold change vs. all for type 1 genes in myositis groups.** Values only included if logFC > 0.

| Gene | DM | | IMNM | | ASyS | | IBM | | PM/Scl | |
|---|---|---|---|---|---|---|---|---|---|---|
| | q-value | logFC | q-value | logFC | q-value | logFC | q-value | logFC | q-value | logFC |
| CCL3 | 3.91E-02 | 0.545 | 9.58E-01 | 0.030 | 1.19E-02 | 0.990 | 1.35E-01 | 0.571 | 1.41E-01 | 0.966 |
| CCL4 | 1.17E-01 | 0.373 | | | 1.49E-02 | 0.861 | 4.98E-07 | 1.426 | 3.13E-02 | 1.145 |
| CCL5 | 7.04E-02 | 0.326 | | | 2.54E-01 | 0.425 | 8.15E-24 | 1.968 | 3.80E-03 | 1.113 |
| CXCL10 | 8.04E-17 | 2.046 | | | 1.43E-02 | 1.105 | 5.42E-07 | 1.788 | 1.46E-02 | 1.591 |
| CXCL11 | 4.11E-35 | 2.804 | | | 3.19E-02 | 1.011 | 3.38E-05 | 1.536 | 5.65E-02 | 1.313 |
| CXCL9 | | | | | 9.85E-07 | 1.759 | 1.36E-21 | 2.951 | 3.48E-06 | 2.573 |
| EBI3 | 4.92E-01 | 0.153 | | | 8.89E-03 | 0.799 | 9.00E-03 | 0.742 | 1.45E-04 | 1.565 |
| IFNG | | | | | 7.42E-02 | 0.857 | 4.61E-18 | 2.601 | 1.38E-02 | 1.501 |
| IL12A | 4.64E-05 | 0.610 | | | 8.34E-01 | 0.116 | 1.91E-02 | 0.525 | 8.04E-01 | 0.149 |
| IL18 | 2.01E-15 | 0.808 | 6.43E-03 | 0.375 | 1.90E-01 | 0.309 | | | | |
| IL27 | 9.14E-10 | 1.180 | | | 8.43E-01 | 0.143 | 4.03E-04 | 1.016 | 7.52E-03 | 1.278 |
| LTA | 7.32E-01 | 0.078 | | | 1.96E-01 | 0.511 | 2.73E-07 | 1.283 | 1.92E-04 | 1.526 |
| TNF | 3.07E-01 | 0.181 | | | 1.70E-01 | 0.448 | 1.88E-06 | 1.027 | 7.95E-03 | 1.002 |
| CCR5 | 9.60E-01 | 0.015 | | | 4.18E-01 | 0.462 | 4.73E-20 | 2.327 | 2.13E-03 | 1.534 |
| CXCR3 | | | | | 6.31E-01 | 0.333 | 7.74E-23 | 2.474 | 5.01E-04 | 1.719 |
| IFNGR1 | 2.23E-09 | 0.293 | 2.88E-04 | 0.220 | 8.78E-01 | 0.029 | | | | |
| IFNGR2 | 4.24E-03 | 0.153 | 2.86E-10 | 0.359 | 9.87E-01 | 0.004 | 2.41E-01 | 0.094 | | |
| IL12RB2 | 8.75E-15 | 0.920 | 3.82E-01 | 0.176 | | | | | 9.91E-01 | 0.008 |
| IL18R1 | 3.90E-06 | 0.491 | | | 6.77E-01 | 0.142 | 2.84E-02 | 0.355 | 9.83E-01 | 0.012 |
| IL18RAP | 1.59E-02 | 0.428 | | | 3.54E-01 | 0.372 | 1.18E-02 | 0.631 | | |
| IL27RA | | | | | 9.83E-01 | 0.008 | 8.09E-03 | 0.334 | 4.49E-04 | 0.676 |
| LTBR | 2.40E-01 | 0.072 | 7.01E-01 | 0.039 | | | | | 2.87E-01 | 0.176 |
| TNFRSF1A | 2.08E-10 | 0.359 | 7.21E-03 | 0.200 | 9.40E-01 | 0.019 | | | 4.76E-01 | 0.138 |
| TNFRSF1B | 2.30E-02 | 0.193 | | | 5.43E-01 | 0.136 | 9.41E-05 | 0.451 | 2.76E-02 | 0.452 |

| Gene | Mi2 | | MDA5 | | NXP2 | | TIF1 | | HMGCR | |
|---|---|---|---|---|---|---|---|---|---|---|
| | q-value | logFC | q-value | logFC | q-value | logFC | q-value | logFC | q-value | logFC |
| CCL3 | 1.88E-02 | 1.290 | 9.96E-01 | 0.014 | | | 7.98E-01 | 0.233 | | |
| CCL4 | 2.66E-01 | 0.568 | 9.73E-01 | 0.082 | 8.90E-01 | 0.176 | 9.13E-01 | 0.099 | | |
| CCL5 | 5.02E-01 | 0.284 | 4.68E-01 | 0.840 | 9.93E-01 | 0.012 | 7.56E-01 | 0.187 | | |
| CXCL10 | 8.61E-03 | 1.600 | 2.98E-01 | 1.658 | 1.64E-04 | 2.040 | 2.63E-02 | 1.429 | | |
| CXCL11 | 2.23E-04 | 2.002 | 1.89E-03 | 2.600 | 4.27E-08 | 2.601 | 1.23E-07 | 2.352 | | |
| CXCL9 | | | | | | | | | | |
| EBI3 | 3.65E-01 | 0.419 | | | | | | | | |
| IFNG | | | | | | | | | | |
| IL12A | 4.37E-02 | 0.696 | | | 1.73E-01 | 0.559 | 9.47E-02 | 0.665 | | |
| IL18 | 2.58E-03 | 0.748 | 9.45E-01 | 0.077 | 5.52E-07 | 1.117 | 2.23E-03 | 0.737 | 1.77E-03 | 0.479 |
| IL27 | 6.97E-04 | 1.513 | 3.46E-01 | 1.228 | 8.32E-02 | 0.873 | 2.77E-01 | 0.670 | | |

| | | | | | | | | | | |
|---|---|---|---|---|---|---|---|---|---|---|
| LTA | 4.87E-01 | 0.328 | | | | | 9.30E-01 | 0.070 | | |
| TNF | 5.23E-01 | 0.254 | | | 9.65E-01 | 0.052 | 9.11E-01 | 0.073 | | |
| CCR5 | 7.76E-01 | 0.180 | | | | | | | | |
| CXCR3 | 7.63E-01 | 0.188 | 9.88E-01 | 0.035 | | | | | | |
| IFNGR1 | 4.39E-03 | 0.335 | 6.90E-01 | 0.196 | 1.41E-02 | 0.306 | 1.34E-01 | 0.214 | 1.47E-02 | 0.181 |
| IFNGR2 | 1.92E-02 | 0.285 | | | 6.29E-01 | 0.096 | 4.64E-01 | 0.119 | 7.68E-08 | 0.355 |
| IL12RB2 | 2.37E-02 | 0.684 | 4.22E-01 | 0.699 | 3.30E-06 | 1.225 | 8.76E-02 | 0.588 | 5.38E-01 | 0.158 |
| IL18R1 | 7.66E-02 | 0.437 | | | 2.18E-01 | 0.384 | 3.02E-02 | 0.569 | | |
| IL18RAP | 3.98E-01 | 0.345 | 9.75E-01 | 0.056 | 7.71E-01 | 0.226 | 4.74E-01 | 0.384 | | |
| IL27RA | | | 8.92E-01 | 0.109 | | | | | | |
| LTBR | 7.51E-01 | 0.048 | | | | | 6.41E-01 | 0.087 | 6.28E-01 | 0.057 |
| TNFRSF1A | 6.67E-04 | 0.447 | 4.29E-01 | 0.329 | 4.99E-03 | 0.391 | 2.29E-01 | 0.214 | 1.82E-02 | 0.206 |
| TNFRSF1B | 1.27E-01 | 0.280 | 8.96E-01 | 0.101 | | | 5.53E-01 | 0.155 | | |

| Gene | SRP | | Jo1 | |
|---|---|---|---|---|
| | q-value | logFC | q-value | logFC |
| CCL3 | 9.71E-01 | 0.149 | 1.97E-04 | 1.582 |
| CCL4 | | | 9.16E-05 | 1.457 |
| CCL5 | | | 1.98E-03 | 0.995 |
| CXCL10 | | | 1.71E-06 | 2.096 |
| CXCL11 | | | 1.08E-05 | 1.962 |
| CXCL9 | | | 2.33E-15 | 2.982 |
| EBI3 | | | 1.72E-02 | 0.935 |
| IFNG | | | 3.13E-04 | 1.680 |
| IL12A | | | 5.77E-02 | 0.623 |
| IL18 | 1.00E+00 | 0.000 | 3.64E-02 | 0.509 |
| IL27 | | | 2.80E-01 | 0.537 |
| LTA | | | 5.92E-02 | 0.776 |
| TNF | | | 1.01E-01 | 0.591 |
| CCR5 | | | 3.59E-02 | 0.996 |
| CXCR3 | | | 5.84E-02 | 0.924 |
| IFNGR1 | 1.87E-01 | 0.298 | 4.54E-01 | 0.102 |
| IFNGR2 | 1.70E-01 | 0.306 | 3.24E-01 | 0.130 |
| IL12RB2 | 9.17E-01 | 0.200 | | |
| IL18R1 | | | 1.96E-01 | 0.340 |
| IL18RAP | | | 3.53E-01 | 0.402 |
| IL27RA | | | 8.38E-01 | 0.054 |
| LTBR | | | | |
| TNFRSF1A | 8.08E-01 | 0.146 | 2.72E-01 | 0.166 |
| TNFRSF1B | | | 6.55E-02 | 0.331 |

**Supplementary Table S4. Adjusted p-values (q-values) of differential overexpression and log-fold change vs. all for type 2 genes in myositis groups.** Values only included if logFC > 0.

| Gene | DM | | IMNM | | ASyS | | IBM | | PM/Scl | |
|---|---|---|---|---|---|---|---|---|---|---|
| | q-value | logFC | q-value | logFC | q-value | logFC | q-value | logFC | q-value | logFC |
| AREG | 1.15E-11 | 1.522 | 7.76E-01 | 0.137 | | | 7.71E-01 | 0.127 | | |
| CCL1 | | | | | 4.48E-01 | 0.231 | 4.50E-04 | 0.639 | 5.57E-01 | 0.246 |
| CCL11 | 2.52E-15 | 1.775 | 8.11E-01 | 0.115 | 1.33E-02 | 1.001 | 3.03E-01 | 0.391 | 2.78E-01 | 0.761 |
| CCL17 | | | 3.67E-01 | 0.288 | | | 4.67E-09 | 1.667 | 6.63E-01 | 0.324 |
| CCL22 | | | 9.62E-01 | 0.023 | | | 2.09E-07 | 1.460 | 4.68E-02 | 1.049 |
| CCL24 | | | | | | | 8.88E-03 | 0.768 | 7.11E-01 | 0.273 |
| CCL26 | 9.24E-01 | 0.024 | 5.90E-01 | 0.177 | | | 1.08E-04 | 1.039 | | |
| IL13 | 6.81E-01 | 0.084 | | | | | 2.45E-02 | 0.573 | | |
| IL1A | | | | | 1.62E-04 | 0.877 | 8.30E-03 | 0.591 | 4.87E-01 | 0.353 |
| IL25 | | | 7.53E-01 | 0.080 | | | 4.19E-02 | 0.434 | | |
| IL33 | | | 1.02E-05 | 0.546 | 2.46E-01 | 0.277 | 2.96E-01 | 0.179 | 8.30E-01 | 0.098 |
| IL4 | | | | | | | | | | |
| IL5 | | | | | 7.20E-01 | 0.152 | 1.80E-01 | 0.261 | 1.46E-01 | 0.582 |
| IL9 | | | | | | | | | | |
| TNFSF15 | 1.60E-01 | 0.208 | | | 3.85E-01 | 0.292 | 1.30E-04 | 0.746 | 4.83E-02 | 0.670 |
| TNFSF4 | 2.06E-01 | 0.180 | | | 5.49E-02 | 0.453 | 2.01E-03 | 0.588 | 2.58E-01 | 0.425 |
| TSLP | 4.34E-02 | 0.216 | 8.49E-05 | 0.458 | | | 9.57E-01 | 0.010 | | |
| CCR3 | | | | | | | 2.88E-01 | 0.349 | | |
| CCR4 | | | | | 8.73E-01 | 0.126 | 2.42E-09 | 1.568 | 3.08E-04 | 1.585 |
| CCR8 | | | | | 6.54E-01 | 0.224 | 6.58E-09 | 1.459 | 2.23E-02 | 1.113 |
| CRLF2 | 9.72E-01 | 0.008 | | | 7.81E-01 | 0.170 | 4.54E-01 | 0.205 | 1.35E-02 | 1.153 |
| EGFR | | | 9.83E-01 | 0.004 | | | 7.42E-01 | 0.042 | | |
| IL13RA1 | | | 5.08E-07 | 0.241 | 6.70E-01 | 0.056 | 8.94E-01 | 0.010 | | |
| IL17RB | 9.64E-01 | 0.006 | 9.81E-01 | 0.006 | | | | | | |
| IL1RL1 | 6.14E-08 | 1.029 | | | 5.17E-02 | 0.691 | | | | |
| IL4R | 7.35E-01 | 0.037 | | | 4.06E-01 | 0.186 | 5.03E-02 | 0.269 | 1.51E-03 | 0.673 |
| IL5RA | | | | | | | 1.65E-12 | 1.821 | 2.30E-01 | 0.695 |
| IL9R | | | | | | | 1.17E-02 | 0.740 | | |
| TNFRSF25 | | | | | 2.29E-01 | 0.251 | 1.33E-02 | 0.357 | 1.81E-08 | 1.124 |
| TNFRSF4 | 4.80E-02 | 0.298 | | | 9.68E-01 | 0.025 | | | 1.04E-07 | 1.533 |

| Gene | Mi2 | | MDA5 | | NXP2 | | TIF1 | | HMGCR | |
|---|---|---|---|---|---|---|---|---|---|---|
| | q-value | logFC | q-value | logFC | q-value | logFC | q-value | logFC | q-value | logFC |
| AREG | 4.84E-03 | 1.485 | 9.03E-01 | 0.302 | 1.93E-03 | 1.603 | 5.32E-03 | 1.444 | 9.49E-01 | 0.048 |
| CCL1 | | | | | | | | | | |
| CCL11 | 4.17E-05 | 1.994 | 8.80E-01 | 0.393 | 2.82E-02 | 1.264 | 5.75E-03 | 1.494 | 9.62E-01 | 0.036 |
| CCL17 | | | | | | | | | 1.13E-01 | 0.529 |
| CCL22 | | | | | | | | | 7.27E-01 | 0.169 |

| | | | | | | | | | | |
|---|---|---|---|---|---|---|---|---|---|---|
| CCL24 | | | | | | | | | | |
| CCL26 | 1.26E-01 | 0.668 | 7.38E-01 | 0.645 | | | | | 3.88E-01 | 0.297 |
| IL13 | | | 8.08E-01 | 0.493 | 9.02E-01 | 0.122 | 8.35E-01 | 0.134 | | |
| IL1A | | | 8.95E-01 | 0.198 | | | | | | |
| IL25 | | | 9.99E-01 | 0.002 | | | | | 4.72E-01 | 0.191 |
| IL33 | | | | | | | | | 5.68E-04 | 0.504 |
| IL4 | | | | | | | | | | |
| IL5 | | | | | | | | | | |
| IL9 | | | | | | | | | | |
| TNFSF15 | | | 6.57E-01 | 0.543 | 8.29E-01 | 0.144 | 8.61E-01 | 0.092 | | |
| TNFSF4 | 8.56E-01 | 0.067 | | | | | 7.38E-01 | 0.151 | 9.07E-01 | 0.041 |
| TSLP | 4.13E-01 | 0.200 | | | 3.40E-01 | 0.303 | 9.49E-01 | 0.028 | 5.83E-05 | 0.530 |
| CCR3 | | | | | 9.59E-01 | 0.082 | 9.10E-01 | 0.098 | | |
| CCR4 | | | | | 9.62E-01 | 0.073 | | | | |
| CCR8 | | | | | | | | | | |
| CRLF2 | | | 6.06E-01 | 0.854 | 2.99E-01 | 0.585 | | | | |
| EGFR | | | | | | | | | 9.79E-01 | 0.006 |
| IL13RA1 | | | | | | | | | 3.28E-05 | 0.232 |
| IL17RB | 8.23E-01 | 0.067 | 6.60E-01 | 0.443 | 9.48E-01 | 0.050 | | | | |
| IL1RL1 | 5.14E-02 | 0.891 | | | 1.19E-04 | 1.530 | 2.27E-01 | 0.703 | | |
| IL4R | 5.98E-01 | 0.122 | | | | | 7.14E-01 | 0.113 | | |
| IL5RA | | | | | | | | | | |
| IL9R | | | | | | | | | | |
| TNFRSF25 | | | | | | | | | | |
| TNFRSF4 | 2.56E-01 | 0.377 | 7.48E-01 | 0.463 | | | 6.78E-01 | 0.199 | | |

| | **SRP** | | **Jo1** | |
|---|---|---|---|---|
| **Gene** | **q-value** | **logFC** | **q-value** | **logFC** |
| AREG | 9.18E-01 | 0.379 | 5.11E-01 | 0.442 |
| CCL1 | | | 1.04E-01 | 0.449 |
| CCL11 | 9.24E-01 | 0.348 | 2.13E-03 | 1.447 |
| CCL17 | | | | |
| CCL22 | | | | |
| CCL24 | 9.93E-01 | 0.032 | | |
| CCL26 | | | | |
| IL13 | | | | |
| IL1A | | | 3.78E-02 | 0.700 |
| IL25 | | | | |
| IL33 | 2.51E-01 | 0.574 | 1.15E-01 | 0.397 |
| IL4 | | | | |
| IL5 | 9.69E-01 | 0.073 | 8.03E-01 | 0.093 |
| IL9 | | | | |
| TNFSF15 | | | 8.50E-03 | 0.729 |
| TNFSF4 | | | 1.13E-03 | 0.804 |

| | | | | |
|---|---|---|---|---|
| TSLP | 9.11E-01 | 0.162 | | |
| CCR3 | | | | |
| CCR4 | | | 9.18E-01 | 0.072 |
| CCR8 | | | 4.04E-01 | 0.355 |
| CRLF2 | 9.23E-01 | 0.256 | 8.57E-01 | 0.097 |
| EGFR | | | | |
| IL13RA1 | 2.69E-01 | 0.222 | 5.14E-01 | 0.074 |
| IL17RB | 9.53E-01 | 0.105 | | |
| IL1RL1 | 9.89E-01 | 0.053 | 7.37E-05 | 1.363 |
| IL4R | 8.57E-01 | 0.191 | 6.18E-02 | 0.374 |
| IL5RA | | | | |
| IL9R | | | | |
| TNFRSF25 | 9.58E-01 | 0.079 | 1.51E-01 | 0.328 |
| TNFRSF4 | | | | |

**Supplementary Table S5. Adjusted p-values (q-values) of differential overexpression and log-fold change vs. all for type 3 genes in myositis groups.** Values only included if logFC > 0.

| Gene | DM | | IMNM | | ASyS | | IBM | | PM/Scl | |
|---|---|---|---|---|---|---|---|---|---|---|
| | q-value | logFC | q-value | logFC | q-value | logFC | q-value | logFC | q-value | logFC |
| CCL20 | 3.29E-02 | 0.500 | 4.37E-01 | 0.264 | 6.70E-06 | 1.370 | 5.81E-01 | 0.184 | 1.48E-01 | 0.871 |
| IL17A | | | 6.48E-01 | 0.095 | | | 2.79E-02 | 0.406 | 5.36E-01 | 0.262 |
| IL17F | | | 7.78E-01 | 0.055 | 8.26E-01 | 0.082 | 4.80E-03 | 0.449 | 2.44E-01 | 0.390 |
| IL1B | | | 6.40E-02 | 0.495 | 2.05E-03 | 0.937 | 6.75E-01 | 0.144 | 4.64E-01 | 0.471 |
| IL22 | | | 8.24E-01 | 0.060 | 8.10E-01 | 0.121 | 2.95E-01 | 0.224 | 2.09E-01 | 0.576 |
| IL23A | 1.66E-01 | 0.237 | | | 2.27E-01 | 0.412 | 5.94E-02 | 0.444 | 1.66E-01 | 0.594 |
| IL26 | | | | | | | | | | |
| IL6 | 1.79E-01 | 0.374 | 4.91E-01 | 0.290 | 5.59E-01 | 0.421 | | | 5.12E-01 | 0.541 |
| CCR6 | 6.79E-02 | 0.563 | | | 3.68E-01 | 0.621 | 2.33E-01 | 0.529 | 1.62E-01 | 1.060 |
| IL17RC | | | | | | | | | 9.79E-01 | 0.009 |
| IL20RA | | | 4.30E-04 | 0.580 | 8.79E-01 | 0.082 | 1.10E-01 | 0.343 | | |
| IL22RA1 | 1.23E-15 | 1.260 | 3.38E-02 | 0.473 | | | 1.33E-01 | 0.398 | 9.65E-01 | 0.038 |
| IL23R | | | | | | | 1.31E-02 | 0.651 | 1.18E-01 | 0.778 |
| IL6R | | | | | 9.35E-01 | 0.030 | | | | |

| Gene | Mi2 | | MDA5 | | NXP2 | | TIF1 | | HMGCR | |
|---|---|---|---|---|---|---|---|---|---|---|
| | q-value | logFC | q-value | logFC | q-value | logFC | q-value | logFC | q-value | logFC |
| CCL20 | 6.74E-03 | 1.425 | | | 8.61E-01 | 0.201 | 7.97E-01 | 0.201 | | |
| IL17A | | | 9.22E-01 | 0.116 | | | | | 6.05E-01 | 0.127 |
| IL17F | | | | | 9.61E-01 | 0.039 | | | 8.66E-01 | 0.044 |
| IL1B | 5.36E-01 | 0.318 | | | | | | | 1.76E-01 | 0.445 |
| IL22 | | | | | 4.06E-01 | 0.426 | | | 9.20E-01 | 0.039 |

| | | | | | | | | | | |
|---|---|---|---|---|---|---|---|---|---|---|
| IL23A | | | | | 3.00E-01 | 0.479 | 8.38E-01 | 0.120 | 9.17E-01 | 0.045 |
| IL26 | | | | | | | | | | |
| IL6 | 1.19E-01 | 0.869 | 8.88E-01 | 0.351 | 9.85E-01 | 0.039 | 9.50E-01 | 0.071 | 6.36E-01 | 0.256 |
| CCR6 | 6.33E-01 | 0.364 | 3.56E-01 | 1.629 | 5.36E-01 | 0.617 | | | | |
| IL17RC | | | 8.83E-01 | 0.099 | | | 7.61E-01 | 0.069 | | |
| IL20RA | 1.41E-01 | 0.468 | | | | | | | 8.36E-03 | 0.519 |
| IL22RA1 | 1.24E-02 | 0.996 | 3.09E-01 | 1.072 | 9.59E-07 | 1.663 | 4.28E-02 | 0.880 | 4.29E-02 | 0.522 |
| IL23R | | | | | | | | | | |
| IL6R | | | 7.95E-01 | 0.264 | | | | | | |

| Gene | SRP | | Jo1 | |
|---|---|---|---|---|
| | **q-value** | **logFC** | **q-value** | **logFC** |
| CCL20 | 4.29E-01 | 0.957 | 4.89E-06 | 1.786 |
| IL17A | | | 9.59E-01 | 0.020 |
| IL17F | 9.61E-01 | 0.074 | 3.94E-01 | 0.227 |
| IL1B | 7.49E-01 | 0.557 | 1.22E-02 | 1.023 |
| IL22 | 9.58E-01 | 0.109 | 7.44E-01 | 0.130 |
| IL23A | | | 3.31E-02 | 0.723 |
| IL26 | | | | |
| IL6 | 9.29E-01 | 0.340 | 5.50E-02 | 1.031 |
| CCR6 | | | 8.36E-02 | 1.100 |
| IL17RC | | | | |
| IL20RA | 3.47E-01 | 0.655 | 2.65E-01 | 0.395 |
| IL22RA1 | 9.27E-01 | 0.238 | | |
| IL23R | | | | |
| IL6R | | | | |

**Supplementary Figure S1. Venn Diagrams of differentially overexpressed genes.** (A) Venn Diagram of differentially overexpressed genes for the 5 myositis clinical groups (DM, IMNM, ASyS, IBM, PM/Scl). For each clinical group, all genes differentially overexpressed for the clinical group or for any of its autoantibody subgroups (e.g., HMGCR and SRP for IMNM) were included in the diagram. (B) Venn Diagram of differentially overexpressed genes for DM and its autoantibody subgroups (Mi2, MDA5, NXP2, and TIF1), excluding genes differentially overexpressed in any other group.

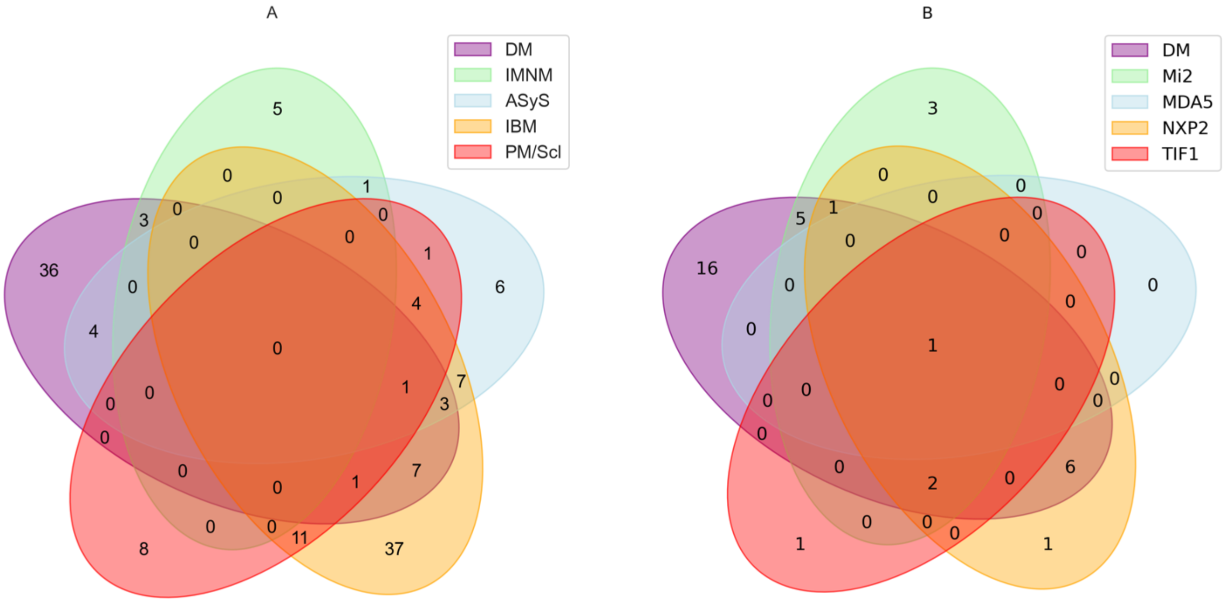

**Supplementary Figure S2. Boxplots of top differentially overexpressed genes by clinical group.** Showing expression of top 2 group-specific differentially overexpressed genes for each clinical group: DM (*TNFSF10*, *GDF15*), IMNM (*IL17B*, *IL13RA1*), ASyS (*CXCL8*, *CCL20*), IBM (*CCL5*, *CCR5*), PM/Scl (*TNFRSF25*, *TNFRSF4*), and NT (*AIMP1*, *MSTN*).

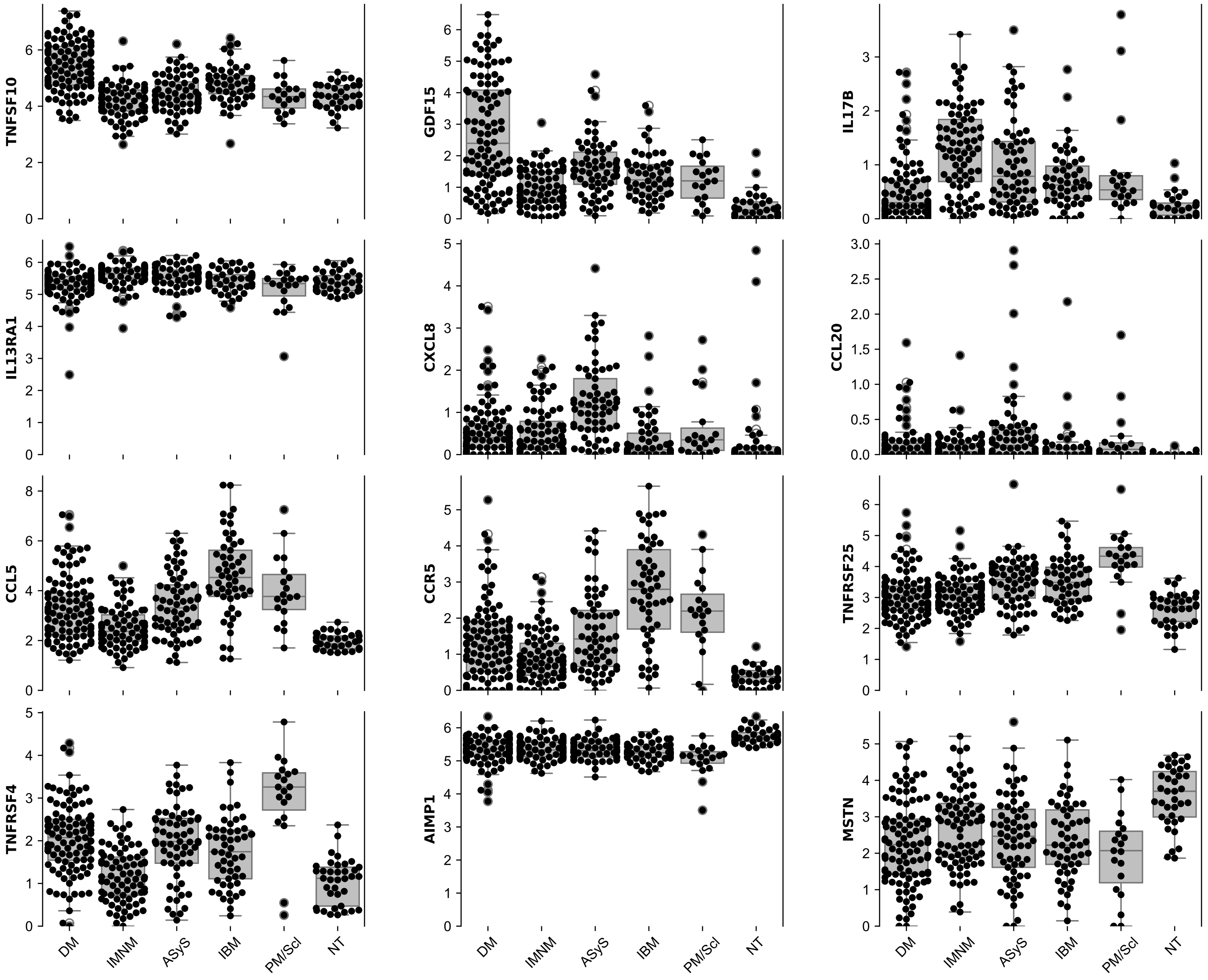

**Supplementary Figure S3. Boxplots of top differentially overexpressed genes by autoantibody subgroup.** Showing expression of top 2 group-specific differentially overexpressed genes for each autoantibody subgroup: Mi2 (*IL11*, *GRN*), NXP2 (*IFNB1*, *IL15RA*), TIF1 (*IL31RA*), HMGCR (*IL17B*, *IL10RB*), Jo1 (CXCL8, *CCL20*). There was only 1 TIF1-specific differentially overexpressed gene, and 0 MDA5- or SRP-specific differentially overexpressed genes.

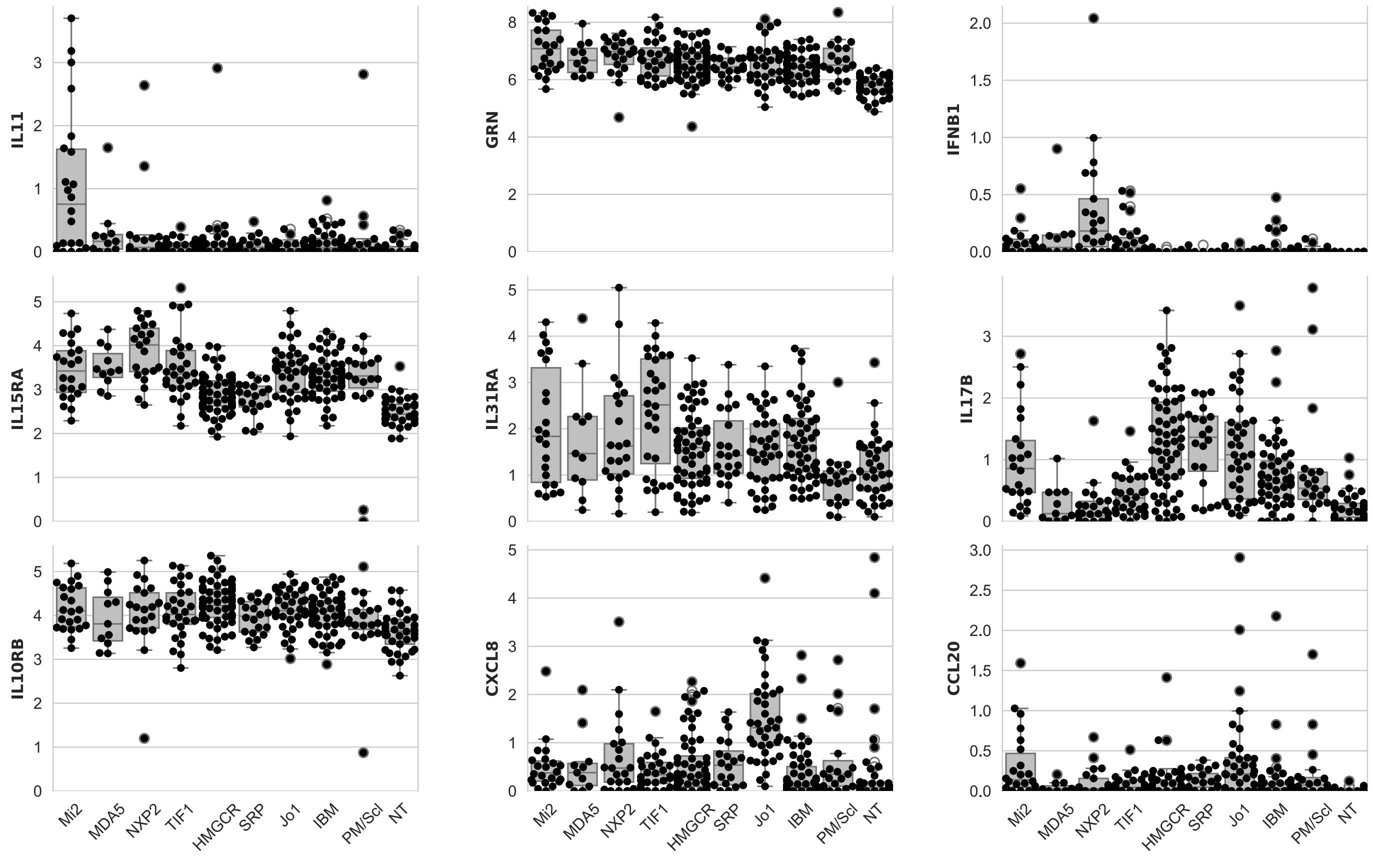
